# One Shot Model For COVID-19 Classification and Lesions Segmentation In Chest CT Scans Using LSTM With Attention Mechanism

**DOI:** 10.1101/2021.02.16.21251754

**Authors:** Aram Ter-Sarkisov

## Abstract

We present a model that fuses instance segmentation, Long Short-Term Memory Network and Attention mechanism to predict COVID-19 and segment chest CT scans. The model works by extracting a sequence of Regions of Interest that contain class-relevant information, and applies two Long Short-Term Memory networks with attention to this sequence to extract class-relevant features. The model is trained in one shot: both segmentation and classification branches, using two different sets of data. We achieve a 95.74% COVID-19 sensitivity, 98.13% Common Pneumonia sensitivity, 99.27% Control sensitivity and 98.15% class-adjusted F1 score on the main dataset of 21191 chest CT scan slices, and also run a number of ablation studies in which we achieve 97.73% COVID-19 sensitivity and 98.41% F1 score. All source code and models are available on https://github.com/AlexTS1980/COVID-LSTM-Attention.

## 1 Introduction

Coronavirus (COVID-19) is an ongoing global pandemic that has taken so far over 91400 lives in the UK alone and over 2.069M worldwide as of late January, 2021 with the crisis worsening in some countries, measured both by the number of deceased and the number of new cases (https://www.worldometers.info/coronavirus). The pandemic caused a complete or partial lockdown in most countries across the planet and led to a previously unseen pressure on healthcare with radiology departments workload exceeding their capacity and manpower.

Analysis of chest CT scans using Deep Learning (DL) can provide assistance to the radiology personnel in many ways. One of them is the reduction of time it takes to process a scan slice from roughly 20 minutes to a few seconds and less [1]. DL algorithms can both rule out clear true positives, and draw the personnel’s attention to suspicious images, e.g. by detecting and segmenting lesions. This may result in two types of errors that the algorithm can possibly make: fail to identify the suspicious areas in scans (false negative) or raise false alarm (false positive) by misclassifying the control image as COVID-19. One of the specific challenges that the personnel, and, therefore, DL algorithms, face is the misclassification of COVID-19 into other types of pneumonia, which is due to a large number of overlaps between the ways these diseases manifest in chest CT scans (see [2–5]).

Existing Deep Learning methodology analyzing chest CT scans has two main limitations: either it relies on large amounts of data (and data manipulation tricks) to train the model or the model was both trained and evaluated on small amounts of data, hence the solution’s ability to extend to larger datasets is debatable. Another problem that, to the best of our knowledge, all DL solutions suffer from, is transferability of results to other datasets without additional finetuning/transfer learning, something that models like Faster R-CNN [6] or Mask R-CNN [7] do not have a problem with due to the training on general-purpose datasets like MS COCO 2017 [8] and Pascal VOC 2012 [9].

One of the approaches in the analysis of images is the extraction of Regions of Interest (RoIs) containing class- or object-specific information. This can be done through either semantic [10] or instance [6, 7] segmentation of objects. In this paper we introduce an advanced model that learns to separate positive/class-relevant and negative/class-irrelevant RoIs using a combination of instance segmentation, Long short-term memory (LSTM) network [11] and attention mechanism [12, 13]. One Shot Attention model leverages Mask R-CNN [7] ability to extract Regions of Interest (RoIs) from images independently to learn the relationships among them for the prediction of the whole image into one of the classes: COVID-19, Commnon Pneumonia (CP) and Control. The novelty and contribution of our work can be summarized in the following way:

1. Advanced architecture that consists of an Attention Layer with two branches independently learning the relationship among class-relevant and class-irrelevant RoIs using LSTM with soft attention,
2. The model is trained and evaluated in one shot, which includes both segmentation and classification branches, only on 3% of the total data, and evaluated on the remaining part thereof,
3. We achieve a 95.35% COVID-19 sensitivity and 98.10% F1 score, which are among the best results for such a large dataset.
4. We quickly extend our results to other datasets using transfer learning and achieve high accuracy.

The rest of the paper is structured in the following way: Section 2 discusses the related literature and the ways we improve on it, Section 3 introduces the data and details of the attention-based methodology, Section 4 discusses experimental setup, results and comparison to the solutions existing in the literature, and also ablation studies. Section 5 concludes.

## 2 Related Work

Mask R-CNN [7] is the state-of-the-art instance segmentation model based on the object detector Faster R-CNN [6]. It predicts the objects’ bounding boxes, classes and masks independently, thus allowing for higher accuracy compared to semantic segmentation model like Fully Convolutional Net (FCN, [10]). The key steps of Mask R-CNN are Region Proposal Net (RPN) that predicts bounding boxes and objects and Region of Interest (RoI) that refines bounding boxes, predicts classes and object masks, see [7] for the details. Mask R-CNN has therefore two branches in RoI: box and mask. We refer to this as segmentation branch. RoI was augmented in [14, 15] to include a classification branch that also has two parallel branches, box and mask: box for the prediction of boxes and mask to extract mask features. Their architecture is identical to the segmentation branch. Weights from the detection and mask segmentation branches are copied into the classification branch during the training phase, which enables the classification branch to detect boxes and extract RoIs from the input image. This provides the classification branch with the following functionality:

1. Detect RoI boxes in any image.
2. These box coordinates are used to resize and crop the corresponding areas in the FPN layer using RoIAlign tool to extract RoI mask features.

Further classification analysis in [15] was built around using these RoI mask features to predict the image class.

Long short-term memory network (LSTM, [11]) is one of the most popular recurrent neural networks (RNNs) used to analyze and extract features from sequential data. The main component of LSTM is a memory cell equipped with three sets of gates (input, forget and output) that control, resp. how much to keep of current information, past information and how much of current information to release. This resolves to a great extent the vanishing gradient problem and enables a more efficient sequence analysis than many other RNNs. With application to COVID-19 diagnosis, in [16] a combined convolutional neural net (ConvNet) and LSTM was presented, in which LSTM takes the last features output of ConvNet (size 512 × 7 × 7) and uses it as an input, and the final fully connected layer connected to LSTM predicts the class of the image (COVID-19, Common Pneumonia and Control).

Attention is one of the most active research topics in deep learning. It was introduced in [13] for machine translation encoder-decoder framework in two versions: soft attention that connects each decoder to all states (weighted average) and hard attention (connection to a single state selected using the alignment score) and [12] in the form of global (connection to all encoder states) and local (connection to a window). [17] introduced a transformer model that replaces recurrent connections such as LSTM with multihead attention embedded in an encoder-decoder framework. It has gained a particular recognition in the natural language processing (NLP) community, e.g. for sentiment classification [18].

There is a number of well-received publications that use a form of attention for COVID-19 prediction and lesion segmentation. In [19] a model with residual connections and attention-aware units was used to predict COVID-19 vs Negative. In [20] attention is computed between convolution maps from two different branches of the model: 2- and 3-class problem classification branches. In [21] attention was used to construct the relationship among separate feature maps extracted by VGG16 [22] from x-ray images.

We improve this approach by replacing whole feature maps with RoI mask features extracted from FPN. This yields better results because irrelevant areas in features maps are ignored; instead, the model focuses on RoIs relevant to the class prediction. Sequence of RoIs is an input in LSTM. Attention mechanism is constructed between an output of LSTM and LSTM’s hidden states.

## 3 Methodology

### 3.1 Data

As in [14, 15, 23, 24] we use CNCB-NCOV dataset and COVIDx-CT splits [24] except that our training data has only 3000 observations (1000/class). Test split with 21191 was used in full. For the segmentation problem, 650 images were used to train and validate the model and 100 for testing. Further dataset details are presented in [15]. The labels for the segmentation problem are derived from the ground truth masks (pixel-level labelling) for 4 classes: background, Ground Glass Opacity (GGO), Consolidation (C) and clean lungs. The last 3 classes are deemed ‘positive’ (i.e. in the sense they are not the background) and each instance thereof has 3 parameters: class label, bounding box coordinates and ground truth mask, each used for model training and validation. This data is used to train the full segmentation branch of the model. Images labelled at a global (image) level are used to train the full classification branch (except RoI classification branch, as explained above).

### 3.2 Model

Overview of the model is presented in Figure 1. Sections 3.3-3.7 describe each component of the model. The main stages of the model are:

**Figure 1:**
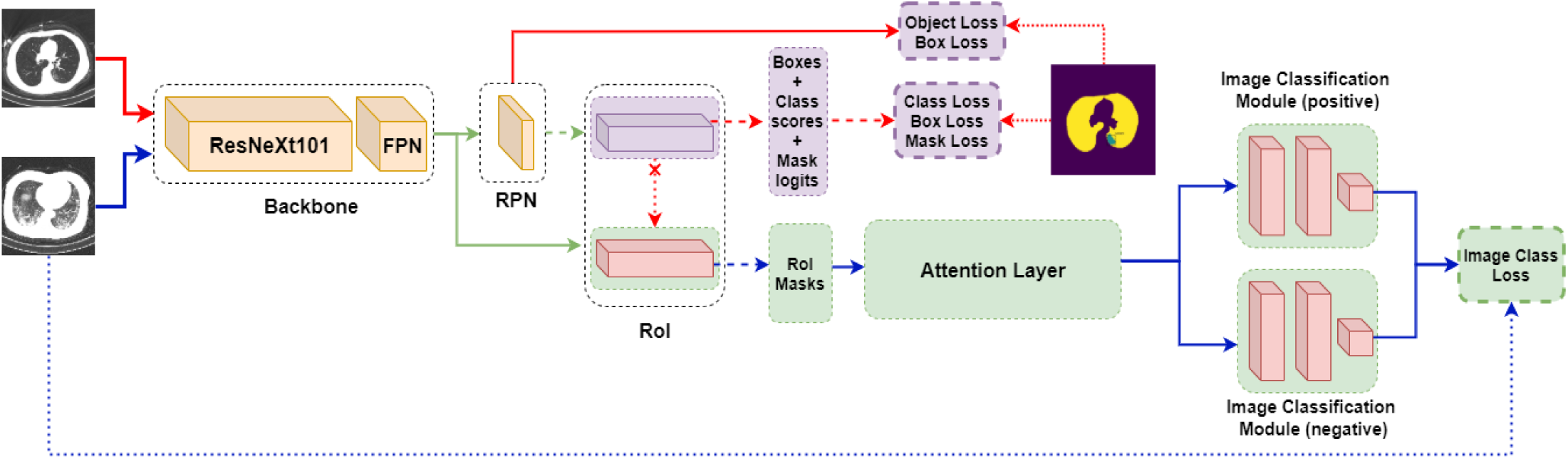
One Shot Model with the Attention Layer (see Figure 2 for its details). Normal arrows: data, Broken arrows: batches or samples, dotted arrows: labels. RoI layer is shared between the segmentation (top) and classification (bottom) branches. Weights are copied from the RoI layer segmentation into the classification branch (red dotted arrow+cross). Full segmentation branch in purple, full classification branch in green. Best viewed in color.

1. Backbone feature extractor,
2. Region Proposal Network (RPN, [7]),
3. Augmented Region of Interest [7, 15] with segmentation and classification branches,
4. Attention layer,
5. Positive (class-relevant) and negative (class-irrelevant) image classification modules,

We refer to the RoI segmentation branch and the corresponding losses as the full segmentation branch and RoI classification branch, Attention layer, image classification modules and the corresponding loss as the full classification branch. Backbone and RPN are shared between the two.

### 3.3 Feature extraction

In this stage a state-of-the-art network with Feature Pyramid Net (FPN) extracts features from the input image. In our setup only the last layer of the backbone, FPN output, is used in RPN layer for box prediction and RoI layer for the RoI feature extraction.

### 3.4 RPN

This layer is inherited from Mask R-CNN. RPN predicts positive (contain object) bounding boxes and passes them to RoI layer. This layer computes loss (bounding box coordinates and object vs background class).

### 3.5 Region of Interest (Segmentation branch)

Our RoI concept is different from [7], as it consists of a total of four branches vs two. Each branch uses RPN box predictions and RoIAlign tool to crop the corresponding area in the FPN feature layer to the predefined size (see [6, 7] for the details). The first two branches, detection branch that predicts boxes and classes and mask segmentation branch are trainable and solve the segmentation problem using object-level labels (bounding boxes, class labels and masks). The remaining two constitute a classification branch and are described in Section 3.6. Segmentation branch is identical to the one in [7].

### 3.6 Region of Interest (Classification branch)

RoI classification branch consists of two branches that have an architecture identical to the ones in the segmentation branch. They are not trainable, instead, they copy weights from the segmentation branch if there is an improvement in the segmentation loss. This lets them detect RoIs and extract mask features size from any image. They do not compute any loss, because their job is to construct and output a batch of RoI mask features size *β*, see [15, 23] for the details. Box predictions are ranked in the order of decreasing confidence score. Since *β* is fixed, the batch size must be the same regardless of the distribution of the confidence scores. This is achieved by setting the confidence threshold *θ* = *−*0.01 to accept even very low-ranking (insignificant) boxes. From these boxes and FPN layer, RoI mask features of fixed size *β × C × H × W* are extracted. Here *β, H, W* are hyperparameters and *C* is the number of feature maps in FPN, also a hyperparameter.

### 3.7 Attention Layer

This is the key step and the novelty of our model. Attention layer consists of two main stages: RoI mask features filtering and LSTM with attention, see Figure 2.

**Figure 2:**
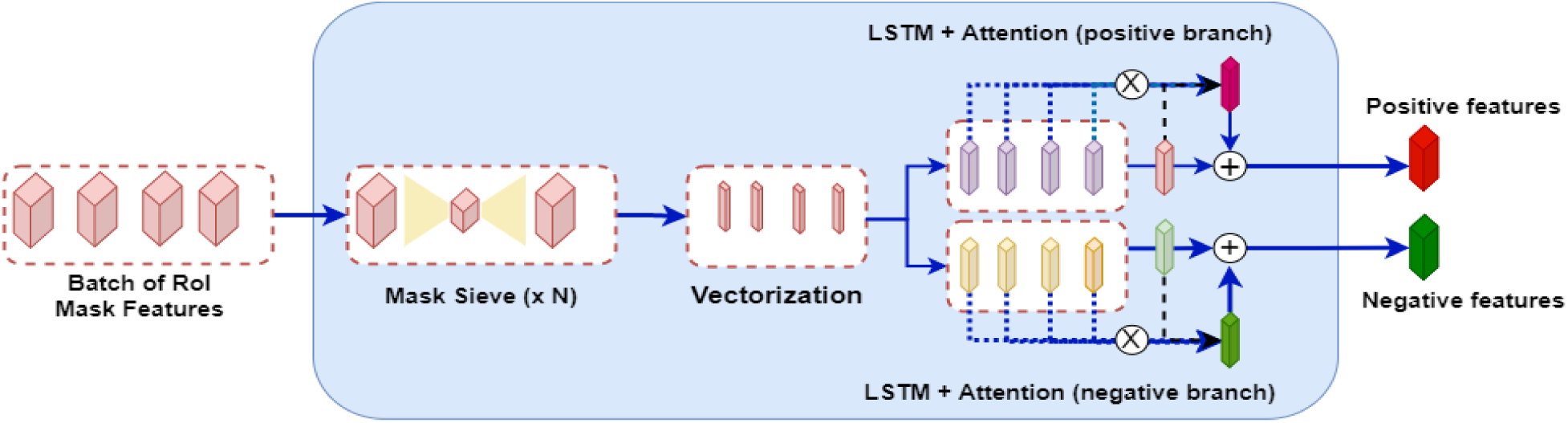
Attention Layer. Features are split into positive (relevant to the class) and negative (irrlevant). Features are fed through LSTM, and attention (dot-product) is computed between the linearized (fully connected) output of LSTM (light red or green) and all LSTM hidden layers. The final output (dark red or dark green) is obtained after the elementwise summation between the attention output (scarlet or green) and the linear layer. For the detailed Attention mechanism see Figure 3. Best viewed in color.

#### 3.7.1 Features filtering

In this stage a small subnetwork filters the batch of RoI mask features to prepare them for attention computation. RoIs in the batch are stored in the decreasing order of their class confidence, and we maintain this order. The rationale for that is that this is an ordered sequence (see Section 4 for the discussion of the order), and LSTM learns sequential relationships. We filter the features through a Mask sieve that first halves the number of feature maps and downsamples their dimensionality using a convolution layer and then upsizes and upsamples them using transposed convolution layer each RoI a total of *N* times: *N ×* (*Conv*2*D → BatchNorm*2*d → Conv*2*DTranspose → ReLU*), therefore the output size stays the same, *β × C × H × W*. This output is vectorized in 3 steps using convolution layer with kernels size 2 × 2, 2 × 2, 7 × 7, such that the output size of the third convolution layer is *β × C*. Finally, before the LSTM input we convert batch dimension *β* to a sequence dimension, keeping the order of the vectorized mask features: 1 *× β × C*.

#### 3.7.2 LSTM+Attention

The detailed architecture of the attention mechanism is presented in Figure 3. We use a form of soft attention [12] that considers all hidden states in LSTM, **H**_**t**_. The main idea of the attention mechanism is to compute a score measuring the relationship between vector **z**_**t**_ that expresses the state of the model after the last LSTM state passed through the linear filter and each hidden LSTM state **h**_*k*_. Unlike [12, 13], we do not have decoder states, as there is no output sequence, therefore the attention is computed for **z**_**t**_. After softmax rescaling, context vector **c**_*t*_ captures the relevant information from the sequence of the hidden states of LSTM.

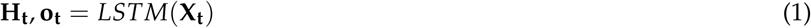

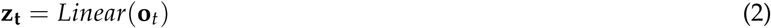

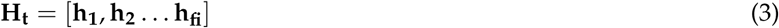

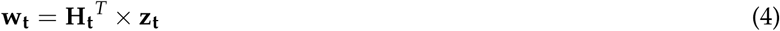

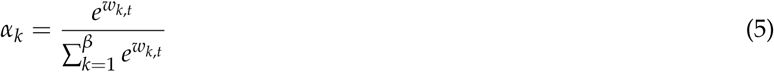

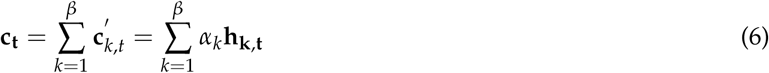

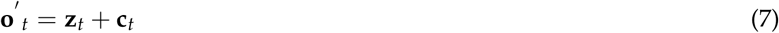

**Figure 3:**
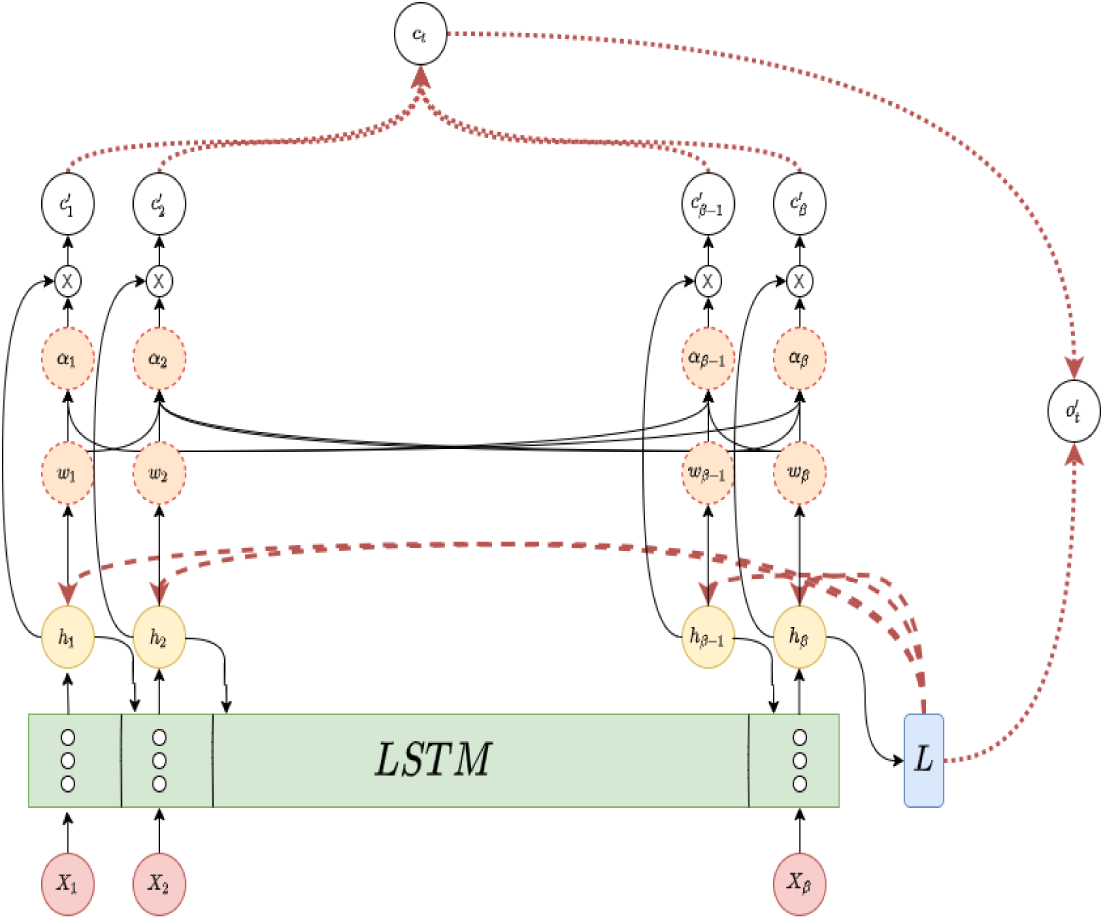
LSTM with Attention Mechanism. *L*: linear layer, broken arrows: dot-product, normal circles: vectors, broken circles: scalars, ⊗:vector-scalar product, dotted arrows: elementwise summation, normal arrows: all other operations. For the explanation of notation see Equations 1-7, subscript *t* omitted. Best viewed in color.

Equation 1 is the LSTM model that processes the RoI batch **X**_**t**_ and outputs the full history **H**_*t*_ (stack of hidden layers, Equation 3) and the last hidden layer **o**_**t**_. Dot-product **w**_**t**_ is rescaled using softmax function (Equations 4, 5). These weights *α*_*k*_ are used to weigh the stack of hidden layers to get the context vector *c*_*t*_, Equation 6. Finally, Equation 7 computes the elementwise sum between the linear vector *z*_*t*_ (Equation 2) and the context vector.

We want to separate out the learning of class-relevant and class-irrelevant RoIs (regardless of their confidence score), and therefore we create two LSTM+Attention: one for the class-relevant (class-positive) RoIs, the other for class-negative RoIs. Their architecture is identical. The key idea of this separation is to prevent the class-positive LSTM from the alignment with the same features regardless of the class. Therefore, Attention layer outputs two sets of linear features: class-positive 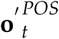 and class-negative 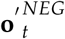.

### 3.8 Image Classification Module and Loss Functions

The architecture of the image classification module is quite simple: two fully connected layers and the output class logits layer with 3 neurons (one/class). The module accepts a vector of features from the Attention layer as an input. As discussed above, the Attention layer has two outputs, 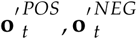, that become inputs in two different modules.

The model solves a segmentation and classification problem simultaneously using a linear combination of two loss functions, Equation 8. Segmentation loss *L*_*SEG*_ is the same as in [7] with three sets of labels: box coordinates, class labels and masks and 5 loss functions: box+object in RPN, box+class+mask in RoI.

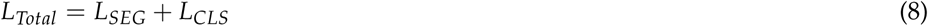

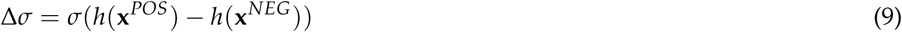

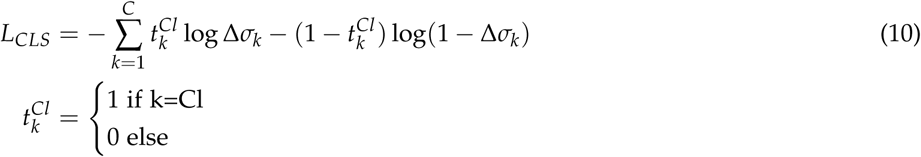

To construct the difference Δ*σ* in Equation 9, the first image classification module learns to return positive class logits *h*(**x**^*POS*^): positive values for the correct image class and negative logits for the incorrect classes and the second image classification module *h*(**x**^*NEG*^) learns to do the opposite: positive values for the incorrect classes and negative value for the correct one. We take the logits difference between the two in Equation 9 so that the expression *h*(**x**^*POS*^) *− h*(**x**^*NEG*^) returns positive values for the correct class and negative for the incorrect ones. *σ* is a sigmoid function computed per-class (class difference in this case). Equation 10 is per-class binary cross-entropy that computes losses for all 3 classes, so we pose the final image classification problem as a multilabel problem with 1 correct and 2 incorrect classes. The total number of classes in our problem is 3: COVID-19, Common Pneumonia and Control/Negative.

## 4 Experimental results

### 4.1 Implementation details

We trained the model using ResNeXt101+Feature Pyramid Network (FPN) [30, 31] with 32 groups (width) each with 8 feature maps (depth) as a feature extractor. The main difference between ResNet101 and ResNeXt101 is the width of the bottleneck module: instead of a single convolution layer, ResNeXt101 has 32 parallel layers (groups) with 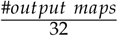 feature maps each connected only to the same number of the corresponding feature maps in the input layer. For example, instead of 256 output maps ResNeXt101 will have 32 layers with 8 feature maps (depth) each. This drastically reduces the number of weights (kernel size = 1 × 1) from 2^16^ = 256 × 256 to 32 × 8 × 8 = 256 × 8 = 2^11^.

The model was trained using Adam [32] optimizer with *β*_1_ = 0.9, *β*_2_ = 0.999, learning rate 1*e −* 5 and weight decay factor 1*e −* 3 for 50 epochs. We compare the trained model to some of the best available in literature. The challenge of comparing results stem from a number of factors:

1. Problem scope: the number of classes does not match, as either two (e.g. COVID-19 vs Control) or three (COVID-19 vs CP vs Control) are reported in the publication,
2. Size of the dataset: very good results (e.g. 99% accuracy) could be reported on very small datasets, which does not necessarily mean that the model could achieve equally good results on much larger datasets,
3. Reported metrics. Some publication report AUC/ROC (area under curve, receiver operating characteristics) instead of F_1_ score, overall accuracy and precision/recall. Not all publications report per-class results.

As a result, this limits the number of publicly available benchmarks to which we can compare our results. Therefore, we attempted two different architectures and compared them to COVID-CT-Mask-Net [23] and a large suite of ResNet and DenseNet [33] models trained on our data. The reported models are:

1. One Shot Model [15]. The model uses a feature Affinity mechanism and also does segmentation+classification in a single shot. Batch size *β* is set to 16, the number of affinities to 8 (see [15] for further details).
2. One Shot Model + LSTM with Attention. The model has the LSTM+attention layer discussed in Section 3.7. LSTM has 256 hidden cells and *β* = 16. Linear layer output **z**_**t**_ has 256 features, matching the hidden dimensions of LSTM. This is necessary for the computation of dot-product alignment in Equation 5.

As discussed above, LSTM uses a sequential input. First, RoIs output from RoI classification branch are ordered in the order of decreasing confidence score, but since one of our objectives is to compute the attention between spatial features, this means that RoIs with different scores could be next to each other in the spatially-aware sequence. Therefore, empirically we found that reordering the sequence of RoIs based on the distance from the origin gives LSTM a better sequential input than confidence scores.

### 4.2 Segmentation Results

We use MS COCO 2017 main criterion [8] in addition to two Intersect over Union (IoU) thresholds: 50% and 75%. To compute the overlap, dot-product between the predicted and ground truth masks is computed. For Average Precision (AP) computation we use Pascal VOC 2012 interpolation: precision is padded with 1 at the start and 0 at the end, recalls are padded with 0 and 1 respectively. Results in Table 1 show that our model outperforms all other architectures trained on CNCB-NCOV data and achieves results comparable to the top-25 of MS COCO leaderboard, mean average precision of 0.470.

**Table 1:** Average Precision on the segmentation test split (100 images). Best results in bold.

| Model | AP@0.5 IoU | AP@0.75 IoU | AP@[0.5:0.95]IoU |
| --- | --- | --- | --- |
| One Shot Model + LSTM+Attention Model | **0.605** | **0.4979** | **0.470** |
| One Shot Model [15] | 0.590 | 0.389 | 0.424 |
| Mask R-CNN | 0.502 | 0.419 | 0.387 |
| Mask R-CNN (heads only) | 0.444 | 0.379 | 0.435 |

#### 4.3 Classification Results

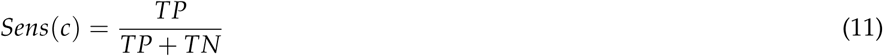

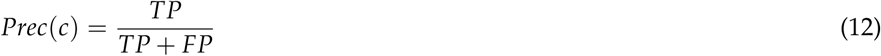

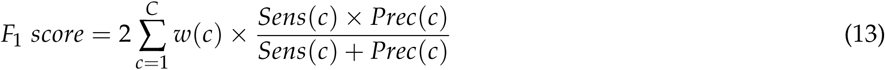

here *w*(*c*) is the class share in the data: *w*(*Neg*) = 45%, *w*(*CP*) = 35%, *w*(*COVID*) = 20%.

Accuracy of the model is computed using sensitivity/recall (Equation 11) and precision/positive predictive value (Equation 12) both for each class and the overall model and *F*_1_ score for the overall model (Equation 13). Sensitivity is the measure of how well the model captures the positive effect given that it exists (false negatives are missed positives), precision is the measure of the ability of the model to avoid finding the effect where it does not exist (false positives). In our implementation of F_1_ score, the weights (shares) of classes in the test set are taken into consideration, to avoid the imbalanced results.

Results in Table 2 show that the model with LSTM+attention outperforms One Shot model with Affinity by about 3% overall, and in COVID-19 sensitivity by about 2.4%. Compared to a suite of baseline ResNet and DenseNet models, LSTM with attention improves the COVID-19 sensitivity by 0.4-7.4% and class-adjusted F_1_ score by 0.2-11%. Considering that a single percentage point may correspond to a large number of human life (lower sensitivity means a larger number of False Negatives), this is a very important and substantial difference.

**Table 2:** Accuracy results of Attention-based and Attention-free models on the COVIDx-CT test split (21191 images). Per-class sensitivity and overall model accuracy are reported. Best results in bold. Results for the reference models were taken from the respective publications and were trained and evaluated on different datasets. COVID-Net CT-2, DenseNet and ResNet models were trained and evaluated on the same data as our model.

| Model | COVID-19 | CP | Negative | F<sub>1</sub>score | Sensitivity | Precision | Dataset |
| --- | --- | --- | --- | --- | --- | --- | --- |
| One Shot Model + LSTM+Attention | **95.74%** | **98.13%** | **99.27%** | **98.15%** | **98.15%** | **98.16%** | 21191 |
| One Shot Model [15] | 93.35% | 95.56% | 95.63% | 95.16% | 95.10% | 95.18% |  |
| COVID-CT-Mask-Net [23] | 90.80% | 91.62% | 91.10% | 91.50% | - | - | 21191 |
| ResNet18 | 92.59% | 96.25% | 92.03% | 93.61% | 93.62% | 93.56% |  |
| ResNet34 | 88.70% | 96.66% | 99.20% | 96.17% | 96.26% | 95.95% |  |
| ResNet50 | 91.04% | 97.64% | 98.97% | 96.88% | 96.89% | 96.84% |  |
| ResNeXt50 | 91.94% | 88.45% | 84.30% | 87.31% | 87.02% | 87.34% |  |
| ResNeXt101 | 91.58% | 92.13% | 94.02% | 92.86% | 93.11% | 92.59% |  |
| DenseNet121 | 92.64% | 96.16% | 98.98% | 96.69% | 96.15% | 97.01% |  |
| DenseNet169 | 89.37% | 96.78% | 98.12% | 95.86% | 96.11% | 95.55% |  |
| DRE-Net [25] | - | - | - | 94% | 93% | 96% | 1282 |
| COVNet [26] | 90% | 87% | 94% | - | - | - | 456 |
| LightCNN [27] | - | - | - | 86.20% | 87.55% | 81.95% | 203 |
| ResNet50 [28] | - | - | - | 74.60% | - | - | 341 |
| DenseNet169 [28] | - | - | - | 76.00% | - | - |  |
| COVID-Net CT-2 [29] | 95.70% | 98.10% | 98.90% | 97.95% | 97.98% | 97.91% | 21191 |

### 4.4 Ablation studies

To establish the ability of the model to further generalize to the unseen data, we perform ablation study on two additional datasets:

1. Hold-out CNCB-NCOV dataset. This is part of the COVIDx-CT train split that we did not use, a total of 58737 images, approx. Negative: 46%, CP: 36% and COVID-19: 18%.
2. iCTCF [34] dataset with 2 classes (Negative and COVID-19), from which we use 600 images (300/class) for training and validation and the remaining 12976 (COVID-19: 9275, Negative: 3701) for testing.

All model hyperparameters were kept the same. Overall results in Tables 3 and 4 confirm the main findings: LSTM with attention outperforms One Shot model with Affinity by about 3% (F_1_ score) and 0.4% (COVID-19 sensitivity) in the CNCB-NCOV holdout set and 1.7% (F_1_ score) and 2.9% (COVID-19 sensitivity) in the iCTCF test set.

**Table 3:** Accuracy results on the COVIDx-CT hold-out split (58737 images). Per-class sensitivity and F1 score are reported. Best results in bold.

| Model | COVID-19 | CP | Negative | F<sub>1</sub>score |
| --- | --- | --- | --- | --- |
| One Shot Model + LSTM+Attention | **98.39%** | **98.28%** | **99.31%** | **98.77%** |
| One Shot Model [15] | 98.02% | 94.15% | 96.09% | 95.75% |

**Table 4:** Accuracy results on the iCTCF-CT test split (12976 images). Per-class sensitivity and F1 score are reported. Best results in bold. Our models were trained with ResNeXt50+FPN backbone.

| Model | COVID-19 | Negative | F<sub>1</sub>score |
| --- | --- | --- | --- |
| One Shot Model + LSTM+Attention | **97.73%** | **98.68%** | **98.41%** |
| One Shot Model | 94.81% | 97.46% | 96.70% |
| VGG16 [34] | 97.00% | 85.47% | - |

## 5 Conclusions

In this paper we presented a novel methodology that combines LSTM with Attention to explore relationship among Regions of Interest feature masks for the purpose of one shot COVID-19 classification and segmentation. The obtained highly accurate results confirm that this method can serve as an assistance diagnostic tool in radiology departments, both to segment instances of lesions and classify whole chest CT scan slices. Our One Shot model with LSTM and attention mechanism achieved 0.470 mean average precision on the test segmentation split, 95.74% COVID-19 precision and 98.15% F_1_ score, some of the best results in the literature on the dataset of this size. Source code of the LSTM with Attention model has been uploaded on https://github.com/AlexTS1980/COVID-LSTM-Attention.

## Data Availability

All data is referenced in the text and is publicly available

## References

[1] Y.-H. Wu, S.-H. Gao, J. Mei, J. Xu, D.-P. Fan, C.-W. Zhao, and M.-M. Cheng, “Jcs: An explainable covid-19 diagnosis system by joint classification and segmentation,” arXiv preprint arXiv:2004.07054, 2020.

[2] D. Zhao, F. Yao, L. Wang, L. Zheng, Y. Gao, J. Ye, F. Guo, H. Zhao, and R. Gao, “A comparative study on the clinical features of covid-19 pneumonia to other pneumonias,” Clinical Infectious Diseases, 2020.

[3] W. Zhao, Z. Zhong, X. Xie, Q. Yu, and J. Liu, “Ct scans of patients with 2019 novel coronavirus (covid-19) pneumonia,” Theranostics, vol. 10, no. 10, p. 4606, 2020.

[4] X. Li, X. Fang, Y. Bian, and J. Lu, “Comparison of chest ct findings between covid-19 pneumonia and other types of viral pneumonia: a two-center retrospective study,” European radiology, pp. 1–9, 2020.

[5] A. Ter-Sarkisov, “Detection and segmentation of lesion areas in chest CT scans for the prediction of COVID-19,” medRxiv, 2020.

[6] S. Ren, K. He, R. Girshick, and J. Sun, “Faster r-cnn: Towards real-time object detection with region proposal networks,” in Advances in neural information processing systems, 2015, pp. 91–99.

[7] K. He, G. Gkioxari, P. Dollár, and R. Girshick, “Mask r-cnn,” in Proceedings of the IEEE international conference on computer vision, 2017, pp. 2961–2969.

[8] T.-Y. Lin, M. Maire, S. Belongie, J. Hays, P. Perona, D. Ramanan, P. Dollár, and C. L. Zitnick, “Microsoft coco: Common objects in context,” in European conference on computer vision. Springer, 2014, pp. 740–755.

[9] M. Everingham, L. Van Gool, C. K. Williams, J. Winn, and A. Zisserman, “The pascal visual object classes (voc) challenge,” International journal of computer vision, vol. 88, no. 2, pp. 303–338, 2010.

[10] J. Long, E. Shelhamer, and T. Darrell, “Fully convolutional networks for semantic segmentation,” in Proceedings of the IEEE conference on computer vision and pattern recognition, 2015, pp. 3431–3440.

[11] S. Hochreiter and J. Schmidhuber, “Long short-term memory,” Neural computation, vol. 9, no. 8, pp. 1735–1780, 1997.

[12] M.-T. Luong, H. Pham, and C. D. Manning, “Effective approaches to attention-based neural machine translation,” arXiv preprint arXiv:1508.04025, 2015.

[13] D. Bahdanau, K. Cho, and Y. Bengio, “Neural machine translation by jointly learning to align and translate,” arXiv preprint arXiv:1409.0473, 2014.

[14] A. Ter-Sarkisov, “Single-shot lightweight model for the detection of lesions and the prediction of covid-19 from chest ct scans,” medRxiv, 2020.

[15] A. Ter-Sarkisov, “One shot model for the prediction of covid-19 and lesions segmentation in chest ct scans through the affinity among lesion mask features,” medRxiv, 2021.

[16] M. Z. Islam, M. M. Islam, and A. Asraf, “A combined deep cnn-lstm network for the detection of novel coronavirus (covid-19) using x-ray images,” Informatics in medicine unlocked, vol. 20, p. 100412, 2020.

[17] A. Vaswani, N. Shazeer, N. Parmar, J. Uszkoreit, L. Jones, A. N. Gomez, L. Kaiser, and I. Polosukhin, “Attention is all you need,” arXiv preprint arXiv:1706.03762, 2017.

[18] J. Zeng, X. Ma, and K. Zhou, “Enhancing attention-based lstm with position context for aspect-level sentiment classification,” IEEE Access, vol. 7, pp. 20462–20471, 2019.

[19] S. Yazdani, S. Minaee, R. Kafieh, N. Saeedizadeh, and M. Sonka, “Covid ct-net: Predicting covid-19 from chest ct images using attentional convolutional network,” arXiv preprint arXiv:2009.05096, 2020.

[20] J. Wang, Y. Bao, Y. Wen, H. Lu, H. Luo, Y. Xiang, X. Li, C. Liu, and D. Qian, “Prior-attention residual learning for more discriminative covid-19 screening in ct images,” IEEE Transactions on Medical Imaging, vol. 39, no. 8, pp. 2572–2583, 2020.

[21] C. Sitaula and M. B. Hossain, “Attention-based vgg-16 model for covid-19 chest x-ray image classification,” Applied Intelligence, pp. 1–14, 2020.

[22] K. Simonyan and A. Zisserman, “Very deep convolutional networks for large-scale image recognition,” arXiv preprint arXiv:1409.1556, 2014.

[23] A. Ter-Sarkisov, “COVID-CT-Mask-Net: Prediction of COVID-19 from CT Scans Using Regional Features,” medRxiv, 2020. [Online]. Available: https://github.com/AlexTS1980/COVID-CT-Mask-Net

[24] H. Gunraj, L. Wang, and A. Wong, “Covidnet-ct: A tailored deep convolutional neural network design for detection of covid-19 cases from chest ct images,” arXiv preprint arXiv:2009.05383, 2020.

[25] Y. Song, S. Zheng, L. Li, X. Zhang, X. Zhang, Z. Huang, J. Chen, H. Zhao, Y. Jie, R. Wang, Y. Chong, J. Shen, Y. Zha, and Y. Yang, “Deep learning enables accurate diagnosis of novel coronavirus (covid-19) with ct images,” medRxiv.

[26] L. Li, L. Qin, Z. Xu, Y. Yin, X. Wang, B. Kong, J. Bai, Y. Lu, Z. Fang, Q. Song et al., “Artificial intelligence distinguishes covid-19 from community acquired pneumonia on chest ct,” Radiology, 2020.

[27] M. Polsinelli, L. Cinque, and G. Placidi, “A light cnn for detecting covid-19 from ct scans of the chest,” arXiv preprint arXiv:2004.12837, 2020.

[28] J. Zhao, Y. Zhang, X. He, and P. Xie, “Covid-ct-dataset: a ct scan dataset about covid-19,” arXiv preprint arXiv:2003.13865, 2020.

[29] H. Gunraj, A. Sabri, D. Koff, and A. Wong, “Covid-net ct-2: Enhanced deep neural networks for detection of covid-19 from chest ct images through bigger, more diverse learning,” 2021.

[30] K. He, X. Zhang, S. Ren, and J. Sun, “Deep residual learning for image recognition,” in Proceedings of the IEEE conference on computer vision and pattern recognition, 2016, pp. 770–778.

[31] T.-Y. Lin, P. Dollár, R. Girshick, K. He, B. Hariharan, and S. Belongie, “Feature pyramid networks for object detection,” in Proceedings of the IEEE conference on computer vision and pattern recognition, 2017, pp. 2117–2125.

[32] D. P. Kingma and J. Ba, “Adam: A method for stochastic optimization,” arXiv preprint arXiv:1412.6980, 2014.

[33] G. Huang, Z. Liu, L. Van Der Maaten, and K. Q. Weinberger, “Densely connected convolutional networks,” in Proceedings of the IEEE conference on computer vision and pattern recognition, 2017, pp. 4700–4708.

[34] W. Ning, S. Lei, J. Yang, Y. Cao, P. Jiang, Q. Yang, J. Zhang, X. Wang, F. Chen, Z. Geng et al., “Open resource of clinical data from patients with pneumonia for the prediction of covid-19 outcomes via deep learning,” Nature biomedical engineering, pp. 1–11, 2020.

